# Entropy-Guided Sample-Specific Feature Selection for Robust Incomplete Multi-Omics Learning in Gut Microbiome Disease Prediction and Biomarker Discovery

**DOI:** 10.1101/2025.10.21.25338061

**Authors:** Min Li, Kaixin Cheng, Mingzhu Lou

## Abstract

The rapid advances in multi-omics data integration technologies have opened unprecedented avenues for dissecting the mechanisms and accelerating the clinical translation of complex diseases. Nevertheless, the frequent absence of certain modalities, coupled with the inherent heterogeneity and high dimensionality of the data, severely restrict the effectiveness of integrative analysis. To address these challenges, we introduce Entropy-guided Sample-Specific Feature Selection for Incomplete Multi-Omics Learning (ESSFS-IMO), a novel framework that couples instance-wise feature selection with entropy-adaptive optimization and variational representation learning. Concretely, ESSFS-IMO leverages a Gumbel– SoftMax selector parameterized by a neural network to achieve per-sample feature selection, while an entropy-based annealing strategy adaptively controls selector sharpness. The selected features are integrated through an information-bottlenecked variational backbone with variance-weighted fusion, enabling robust classification under arbitrary missing patterns. Extensive experiments on inflammatory bowel disease (IBD) multi-omics datasets demonstrate that ESSFS-IMO consistently outperforms state-of-the-art baselines in terms of accuracy, F1 and AUC.

## 1. Introduction

Over the past decade, the gut microbiota has emerged as a central regulator of human health, extending its influence far beyond the digestive tract [1]. Rather than functioning merely as a collection of commensal organisms, the microbiome engages in continuous dialogue with the host immune system, shaping both innate and adaptive responses. These microorganisms also generate a wide array of bioactive metabolites, such as short-chain fatty acids and secondary bile acids, which act as signaling molecules that affect energy balance, endocrine function, and inflammatory pathways [2]. Disturbances in these host–microbe interactions, often triggered by diet, antibiotics, or lifestyle, have been linked to a broad spectrum of conditions ranging from autoimmune and metabolic disorders to cancer and neurological diseases [3][6].

To further unravel how microbial communities influence host physiology, researchers have increasingly turned to advanced omics technologies [7], including metagenomics, metatranscriptomics, and metabolomics. While each of these approaches provides valuable insights into a specific layer of molecular activity or regulatory mechanisms, they remain inherently limited when studied in isolation [8]. Recent investigations suggest that integrating multiple omics layers yields a more comprehensive view of the microbiome–host interplay. Such integrative strategies not only enhance the precision of disease risk assessment but also improve analytical stability and enable the discovery of clinically relevant biomarkers [9][11]. Nevertheless, microbiome-related multi-omics datasets are notoriously complex—characterized by heterogeneity, sparsity, and high dimensionality. These challenges have driven the development of specialized computational frameworks, with machine learning emerging as a particularly powerful paradigm for data integration, feature selection, and predictive modeling. As a result, a growing number of models have been developed to unlock the potential of multi-omics data and advance both biological understanding and clinical applications.

A broad spectrum of computational strategies has been proposed for multi-omics integration, ranging from statistical models and kernel-based approaches to deep learning and graph-based frameworks [12][13]. Among kernel-based methods, Zhang et al. [14] introduced wMKL, a weighted multiple kernel learning framework that integrates diverse omics layers for more accurate cancer subtype identification. Deep learning–based models have also gained traction; for instance, Poirion et al. [15] proposed DeepProg, an ensemble of deep and machine learning models applied to RNA-seq, miRNA, and DNA methylation data for patient risk stratification, while Guo et al. [16] employed a denoising autoencoder framework to integrate mRNA, miRNA, and CNV profiles for ovarian cancer subtyping. More recently, network- and representation-based methods have emerged, such as MDICC by Wang et al. [17], which combines weighted least squares, low-rank subspace representation, and entropy-based fusion for robust subtype discovery, and MVMI by Cano, which leverages ensemble multi-view multi-instance learning to capture complementary information across heterogeneous omics sources. In addition, graph-based models are receiving increasing attention because they can naturally represent multi-omics data as structured networks and enable effective information propagation across heterogeneous feature spaces. For example, MOGONET [18] addresses the heterogeneity among different omics data by leveraging graph convolutional networks (GCN) and view correlation discovery networks (VCDN). These methods effectively integrate multi-omics data and, on this basis, reveal the interactions among different omics data as well as the mechanisms of diseases. Nonetheless, multi-omics data present several intrinsic challenges. First, the dimensionality of such data is often extremely high relative to the number of samples, which easily leads to overfitting; feature selection techniques are sometimes employed to alleviate this issue by filtering redundant variables. Second, incomplete and noisy measurements, resulting from technical limitations or missing modalities, remain prevalent and reduce the robustness of downstream analysis. Third, the strong heterogeneity across different omics layers, together with limited sample size, further complicates integration and hinders model interpretability. [19][20] These issues underscore the need for new methodologies that are both robust to missing information and capable of capturing complementary cross-omics knowledge.

Although many computational frameworks have been proposed to address these challenges, most existing approaches still struggle when confronted with incomplete multi-omics data and often fail to adequately capture sample-specific heterogeneity [21]. Such limitations reduce their effectiveness in disease prediction and biomarker discovery. To address these gaps, we propose Entropy-Guided Sample-Specific Feature Selection for robust Incomplete Multi-Omics Learning (ESSFS-IMO), a novel framework designed to achieve robust and interpretable multi-omics integration under missing-data conditions. The workflow can be summarized in three stages. First, given incomplete multi-omics inputs, the model employs a dynamic Gamma network to generate sample-specific parameters, enabling differentiable feature selection via the Concrete distribution. Second, a confidence-guided adaptive annealing strategy adjusts the temperature according to predictive entropy: samples with high predictive confidence converge rapidly toward sparse selections, whereas those with low confidence preserve exploratory capacity. Finally, the selected features are encoded by modality-specific variational encoders, fused into a joint latent representation through variance-weighted aggregation, and passed to a classifier for prediction. Training is guided by the information bottleneck principle to maximize label-relevant information while minimizing redundant information. In this way, ESSFS-IMO achieves robust learning from incomplete multi-omics datasets while providing interpretable sample-specific feature selection. Extensive experiments on IBD multi-omics datasets further demonstrate that ESSFS-IMO consistently outperforms state-of-the-art baselines in both disease subtype identification and prognostic prediction. The source code of this work can be downloaded from GitHub (https://github.com/KKXXCheng/ESSFS-IMO)

The contributions of this paper are summarized as follows:

1. A novel framework, ESSFS-IMO, is proposed for incomplete multi-omics learning, enabling sample-specific and differentiable feature selection to achieve robust disease prediction.
2. A confidence-aware entropy-guided annealing strategy is introduced, allowing the optimization process to dynamically adapt to predictive uncertainty and thereby enhancing training stability.
3. The framework incorporates a variational neural network backbone that integrates incomplete multiomics data into a compact joint latent representation under the information bottleneck principle
4. Extensive evaluations on multiple real-world multi-omics datasets confirm that ESSFS-IMO surpasses state-of-the-art baselines in predictive accuracy, robustness to missing modalities, and interpretability of selected features.

## 2. Related works

In this section, we first define the incomplete multi-omics learning problem and establish the necessary notation. Then, the overall structure of the proposed ESSFS-IMO framework is introduced, including the entropy-guided sample-specific feature selection module, omics-specific encoding and prediction networks, the joint encoder for modality fusion, and the optimization strategy based on the information bottleneck principle.

### 2.1 Incomplete Multi-Omics Problem Formulation

The emergence of high-throughput sequencing and profiling technologies has enabled the large-scale collection of multi-omics data, allowing researchers to investigate biological systems from multiple complementary perspectives. Integrating data from genomics, transcriptomics, proteomics, metabolomics, and other molecular layers yields a holistic view of complex diseases and facilitates the discovery of clinically relevant biomarkers. In real-world applications, however, multi-omics datasets are frequently incomplete due to sample limitations, financial costs, or technical constraints. The resulting missing modalities substantially hinder the development of integrative predictive models.

The incomplete multi-omics learning problem can be formulated as follows. Consider a dataset contain *N* samples and *V* omics types. For the *n*-th sample, the representation of the *v*-th omics type is denoted by 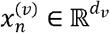,with the corresponding label *y*_*n*_. An indicator variable 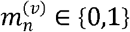 specifies whether this modality is available. The challenge is to construct a mapping 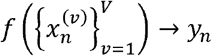 that is robust to arbitrary missing patterns. A model that ignores incomplete cases loses valuable statistical power, while naive imputation risks propagating noise and bias.

For instance, consider a cohort of 800 samples profiled for genomics, transcriptomics, and proteomics. If only 600 samples have complete data across all three omics layers, while the remaining 200 lack one or more omics types, the dataset is deemed incomplete with a missing rate of 25%. Exploiting all 800 samples—rather than restricting analyses to the 600 complete cases—is essential to maximize statistical power and model generalizability. Achieving this without introducing noise or bias remains a central challenge in incomplete multi-omics learning.

### 2.2. Concrete Distribution for Differentiable Feature Selection

A fundamental challenge in multi-omics learning lies in the high dimensionality of omics data, which exhibit redundancy, noise, and a relatively small sample size. Traditional feature selection strategies often rely on discrete choices that are non-differentiable, limiting their compatibility with deep neural networks. To overcome this, the Concrete distribution—also known as the Gumbel–SoftMax distribution—provides a continuous relaxation of categorical variables [22][23].

Given feature logits 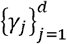. the Concrete distribution perturbs them with Gumbel noise and scales the results by a temperature parameter *T*, yielding the following differentiable approximation:

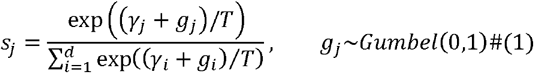

At high values of *T*, the sampling produces smooth probability vectors that encourage exploration across multiple features. As *T* decreases, the distribution becomes increasingly peaked, approaching a one-hot vector and thus effectively selecting individual features. This mechanism allows gradient-based optimization of feature selection processes, enabling end-to-end training. In the context of multi-omics, it provides a principled way to identify compact, discriminative subsets of features while preserving interpretability.

### 2.3. Information Bottleneck Principle in Representation Learning

Beyond feature selection, effective multi-omics integration requires learning latent representations that capture essential predictive information while filtering out noise and redundancy. The Information Bottleneck (IB) principle [24] provides a theoretical foundation for this objective. Specifically, the IB framework encourages the construction of latent variables *Z* that maximize their mutual information with the target label *Y* while minimizing their dependence on the raw input *X*. This trade-off is formally expressed as:

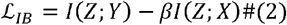

where *β* is a trade-off parameter controlling the balance between relevance and compression.

In multi-omics applications, the IB principle plays two critical roles. First, it reduces the detrimental effect of modality-specific noise [25], which is common in high-dimensional omics data. Second, it ensures that the learned latent representation captures task-relevant signals shared across different omics views. By filtering redundant information and emphasizing predictive sufficiency, IB-based models achieve greater robustness and generalization when facing incomplete or heterogeneous inputs.

### 2.4. Variational Neural Networks for Multi-Omics Integration

While the IB principle defines a conceptual trade-off, variational neural networks (VNNs) [26] provide a practical framework to implement it through variational inference. VNNs combine deep neural architectures with probabilistic latent variable modeling, enabling them to capture both uncertainty and complex structure in multi-omics data. Instead of learning deterministic mappings, VNNs model the conditional distribution over latent variables given the observed input, typically parameterized by mean vector *μ* and variance vector *σ*^2^.

Formally, for each omics input *x*, the encoder network defines an approximate posterior distribution:

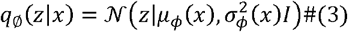

where *ϕ* denotes the encoder parameters. A latent sample is obtained via the reparameterization trick:

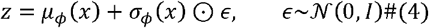

The predictive distribution is then modeled by a decoder or classifier *p*_*θ*_(*y*|*z*), parameterized by *θ*. The training objective maximizes a variational lower bound that combines prediction accuracy with a regularization term:

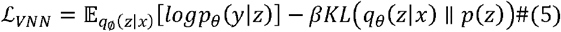

where *p*(*z*) is a prior distribution, often chosen as a standard Gaussian.

In the context of incomplete multi-omics, VNNs offer a principled way to handle missing modalities. Each available omics type contributes to the construction of the latent representation, while absent modalities are naturally ignored in the likelihood term without requiring explicit imputation [27]. This stochastic encoding mechanism allows the model to propagate uncertainty arising from missing data, leading to more robust predictions. Moreover, the shared latent representation facilitates the integration of heterogeneous omics signals, making VNNs well-suited for disease prediction and biomarker discovery where data incompleteness is prevalent.

In summary, prior work has established the theoretical and methodological foundations for addressing incomplete multi-omics learning. Problem formulations highlight the necessity of handling missing modalities without discarding valuable samples. The Concrete distribution introduces a differentiable mechanism for feature selection, while the Information Bottleneck principle provides a rigorous lens for balancing compression with predictive sufficiency. Variational neural networks extend these ideas into a probabilistic deep learning framework that naturally accommodates uncertainty and missing data.

## 3. Methodology

The proposed framework aims to address the challenges of incomplete multi-omics learning by integrating sample-dependent feature selection, adaptive optimization strategies, and probabilistic representation learning. As illustrated in Figure 1, the model comprises three main components: (i) a Dynamic Gamma module for differentiable feature selection, (ii) a confidence-guided adaptive annealing strategy, and (iii) a variational neural network backbone with an information-bottleneck objective. Together, these components form a unified system that ensures robust predictive performance and interpretable feature selection under incomplete data conditions.

**Figure 1.**
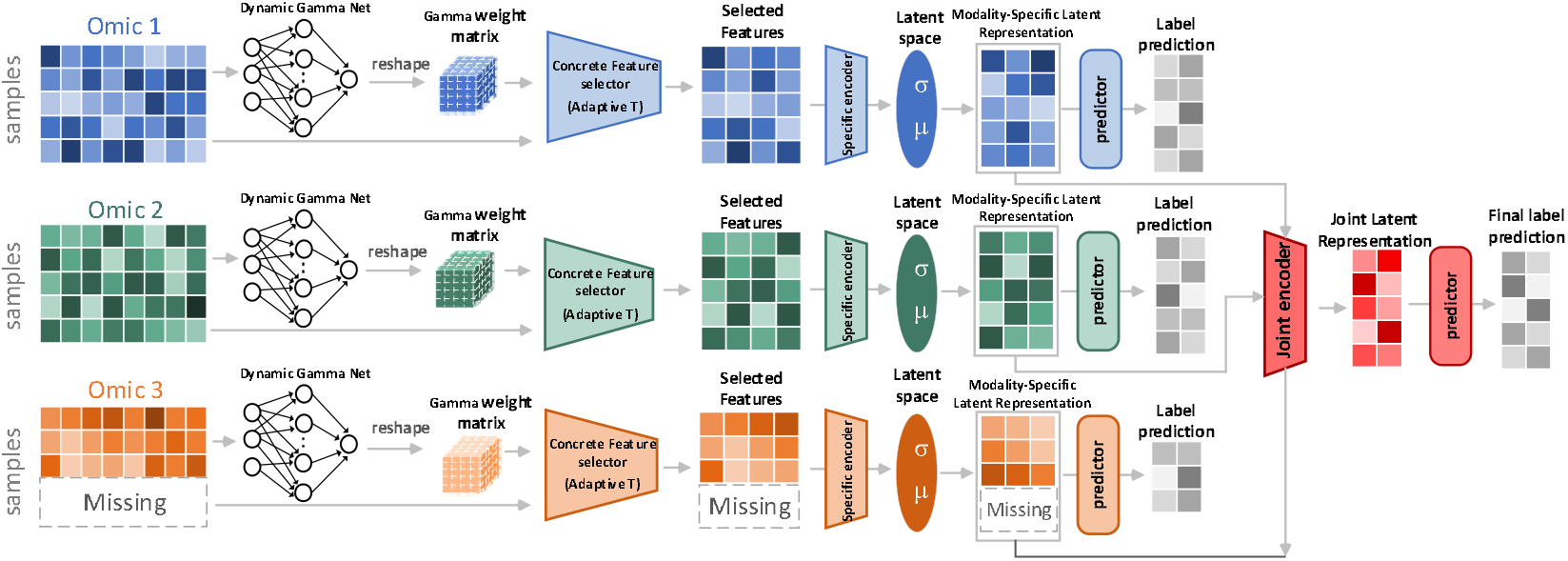
Flowchart of ESSFS-IMO for three omics data. To clearly illustrate the proposed ESSFS-IMO, we assume that the omics3 data of some samples are missing. First, the entropy-guided sample-specific selector adaptively screens out the most discriminative features from each omics data according to the weights output by the dynamic Gamma network. After the screening is completed, the omics-specific encoder maps the selected features to the latent space to obtain the marginal latent representation of the samples. Subsequently, the omics-level predictor uses these marginal latent representations to generate preliminary class inferences. Then, under the constraint of the variational information bottleneck, the joint omics encoder fuses each marginal representation with uncertainty weighting to obtain a joint latent representation. Finally, the joint predictor completes the final classification output based on the joint latent representation and provides support for the interpretation of downstream biomarkers.

### 3.1. Overall Framework of the ESSFS-IMO

The framework begins with multiple omics inputs, each of which may be incomplete due to missing modalities. For each input, a Dynamic Gamma Net generates sample-specific logits that parameterize the Concrete distribution, producing differentiable feature selection vectors. These selected features are then passed through modality-specific encoders, each outputting a latent distribution parameterized by its mean and variance. The encoders use the reparameterization trick to generate stochastic latent embeddings, which are subsequently integrated by a joint variational encoder into a unified latent space. Finally, a classifier operating on this joint representation predicts the target label.

A key innovation is the incorporation of a confidence-guided adaptive annealing mechanism during training, which dynamically adjusts the temperature of the Concrete distribution for each sample. High-confidence samples undergo faster annealing toward discrete feature selections, while low-confidence samples retain smoother distributions to allow further exploration. By combining dynamic feature selection, probabilistic latent variable modeling, and confidence-aware optimization, the framework provides a principled solution to incomplete multi-omics learning.

### 3.2. Dynamic Gamma for Sample-Dependent Concrete Distribution

As described in Section 2.2, the Concrete distribution provides a differentiable approximation for discrete feature selection, enabling end-to-end training of feature selectors within deep neural networks. In prior methods, however, the logits *γ* that parameterize the Concrete distribution are typically treated as static trainable parameters, shared across all samples. While effective for global feature ranking, this static formulation overlooks the heterogeneity among individual samples, which is a critical factor in multi-omics data.

To overcome this limitation, we introduce a dynamic gamma mechanism that adapts feature selection to each input sample. Specifically, instead of optimizing *γ* as a static parameter vector, we design a lightweight neural module—termed the Dynamic Gamma Net—that generates sample-dependent logits conditioned on the raw omics input. Formally, given the omics feature vector 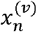, the dynamic logits are defined as:

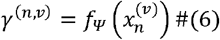

Where *f*_*ψ*_(·) denotes the Dynamic Gamma Net parameterized by *ψ*. These logits parameterize the Concrete distribution with temperature *T*_*n,e*_, yielding a differentiable selection vector:

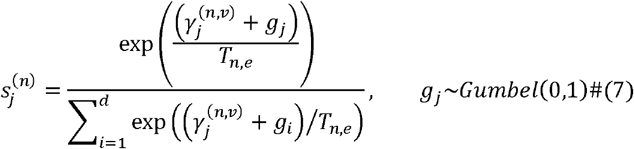

This formulation makes feature selection sample-specific rather than global, enabling the model to adaptively select discriminative features for each sample. Such flexibility is crucial in incomplete multi-omics data, where the predictive signal shift with the available modalities. Moreover, the sample-level selection vectors yield individual-level interpretability, revealing patient-specific drivers of prediction.

### 3.3. Confidence-Guided Adaptive Annealing Strategy

The temperature parameter *T* plays a central role in the Concrete distribution, as it controls the sharpness of the feature selection probabilities. A high temperature encourages exploratory behavior by producing smooth distributions, while a low temperature yields near one-hot vectors that approximate discrete feature selection. Conventional implementations adopt a global exponential annealing schedule, gradually reducing *T* from an initial value *T*_0_ to a final value *T*_*E*_ over training epochs:

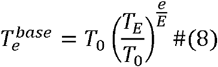

where *e* denotes the current epoch and *E* the total number of epochs. While effective in promoting convergence, this global schedule applies the same cooling rate to all samples, disregarding their varying levels of confidence during learning.

We therefore propose a confidence-guided adaptive annealing strategy, where the temperature is adjusted based on predictive uncertainty. For sample *n*, the entropy of the predictive distribution is:

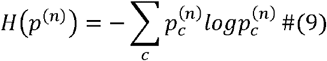

The adaptive temperature is then defined as:

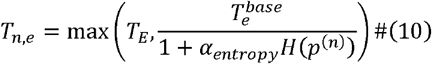

where *α*_*entrophy*_ > 0 controls the sensitivity of the annealing process to entropy. The use *max* (·) ensures that the temperature does not fall below the minimum threshold *T*_*E*_. This formulation results in sample-dependent temperatures: confident samples with low entropy receive lower temperatures earlier, while uncertain samples with high entropy maintain smoother distributions for longer exploration.

By embedding predictive uncertainty into the annealing schedule, the model achieves an improved balance between exploration and exploitation. This strategy is crucial in incomplete multi-omics learning, where the absence of modalities can lead to heterogeneous levels of confidence across samples. Instead of enforcing a uniform training trajectory, the proposed mechanism allows each sample to follow an adaptive path to convergence, ultimately boosting stability and accuracy.

### 3.4. Variational Neural Network Backbone with Information Bottleneck

While the dynamic feature selection and adaptive annealing enhance the model’s capacity to extract relevant inputs, its predictive power hinges on how these features are encoded and fused across modalities. To this end, our architecture employs a variational neural network (VNN) backbone that uses probabilistic inference principles and an information-bottleneck (IB) objective to learn compact, task-relevant latent representations.

For each omics view *v*, the feature-selected representation 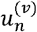 of sample *n* is passed through a modality-specific encoder, yielding the parameters of a latent Gaussian distribution:

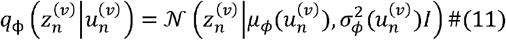

where *μ*_*ϕ*_(·) and *σ*_*ϕ*_(·) are outputs of the encoder network parameterized by *ϕ*. A latent variable is sampled using the reparameterization trick:

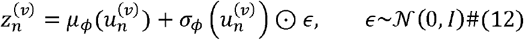

A joint encoder then fuses these modality-specific embeddings into a unified latent representation *Z*_*n*_,using a variance-weighted aggregation strategy that accounts for missing modalities:

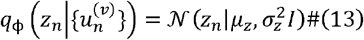

The backbone is trained using a supervised variational objective based on the information bottleneck principle:

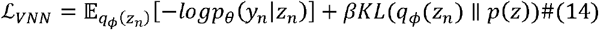

This design ensures that the latent representation retains predictive information while compressing irrelevant redundancy. By explicitly modeling uncertainty, the VNN backbone improves robustness to missing modalities and enhances the biological interpretability of the learned representation.

### 3.5. Training Objective and Optimization

The final training loss integrates the joint supervised variational objective with modality-specific constraints. For each sample *n*, the overall loss is defined as:

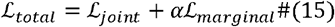

where

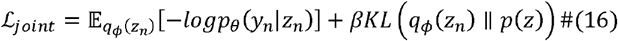

and

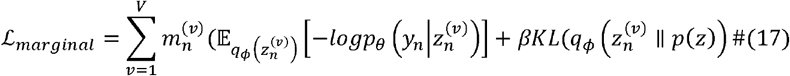

Optimization is carried out using the Adam optimizer, with higher learning rates assigned to the Dynamic Gamma Nets to accelerate the convergence of the gating mechanism. During training, the adaptive temperature *T*_*n,e*_ is updated at each epoch based on predictive entropy, ensuring sample-specific annealing schedules. At inference time, a fixed minimal temperature *T*_*E*_ is applied to obtain sharp feature selections.

The entire procedure is summarized in Algorithm 1, which outlines the interplay between dynamic feature selection, adaptive annealing, and variational inference during training.

#### Algorithm 1

Training procedure of the ESSFS-IMO

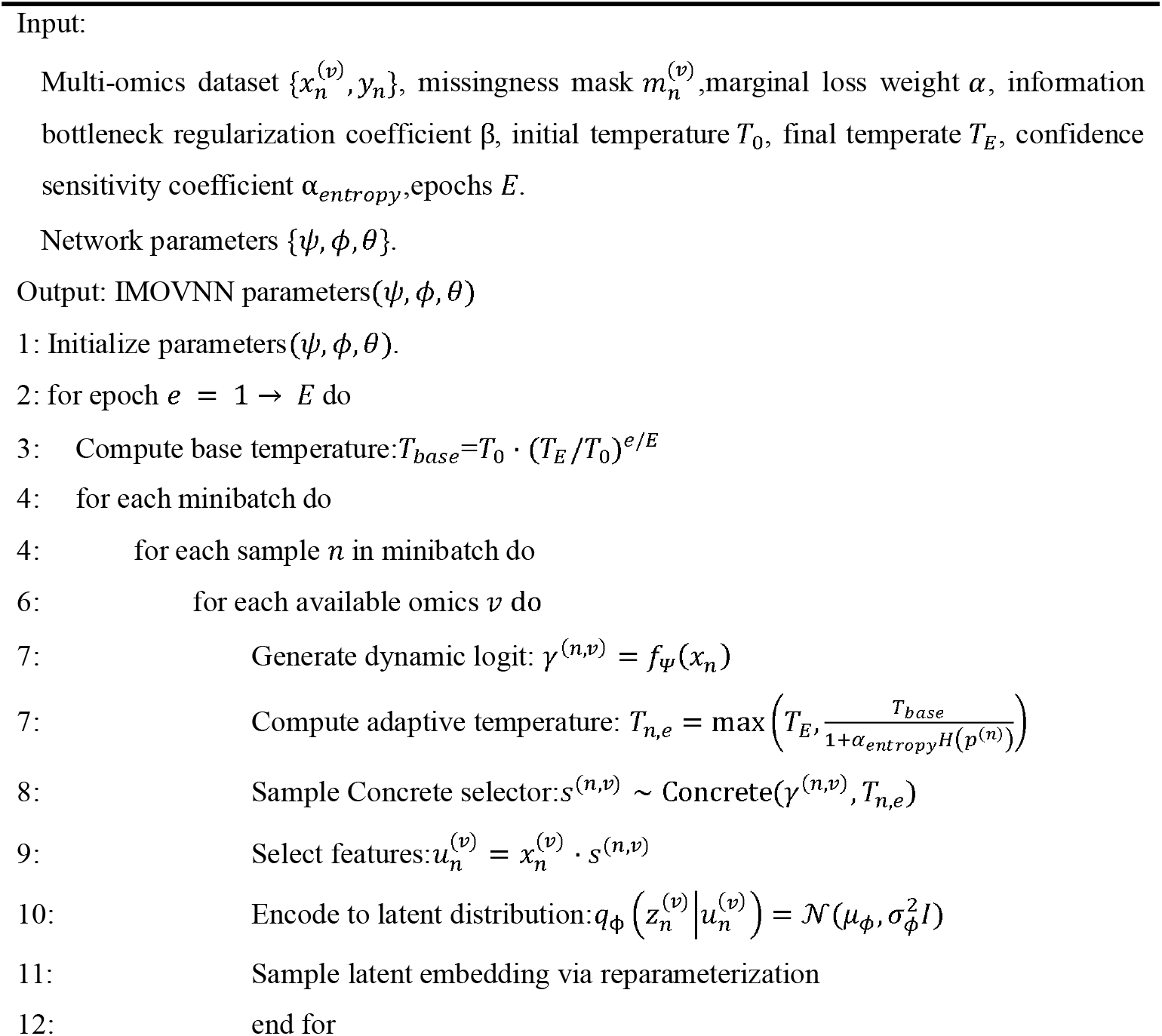
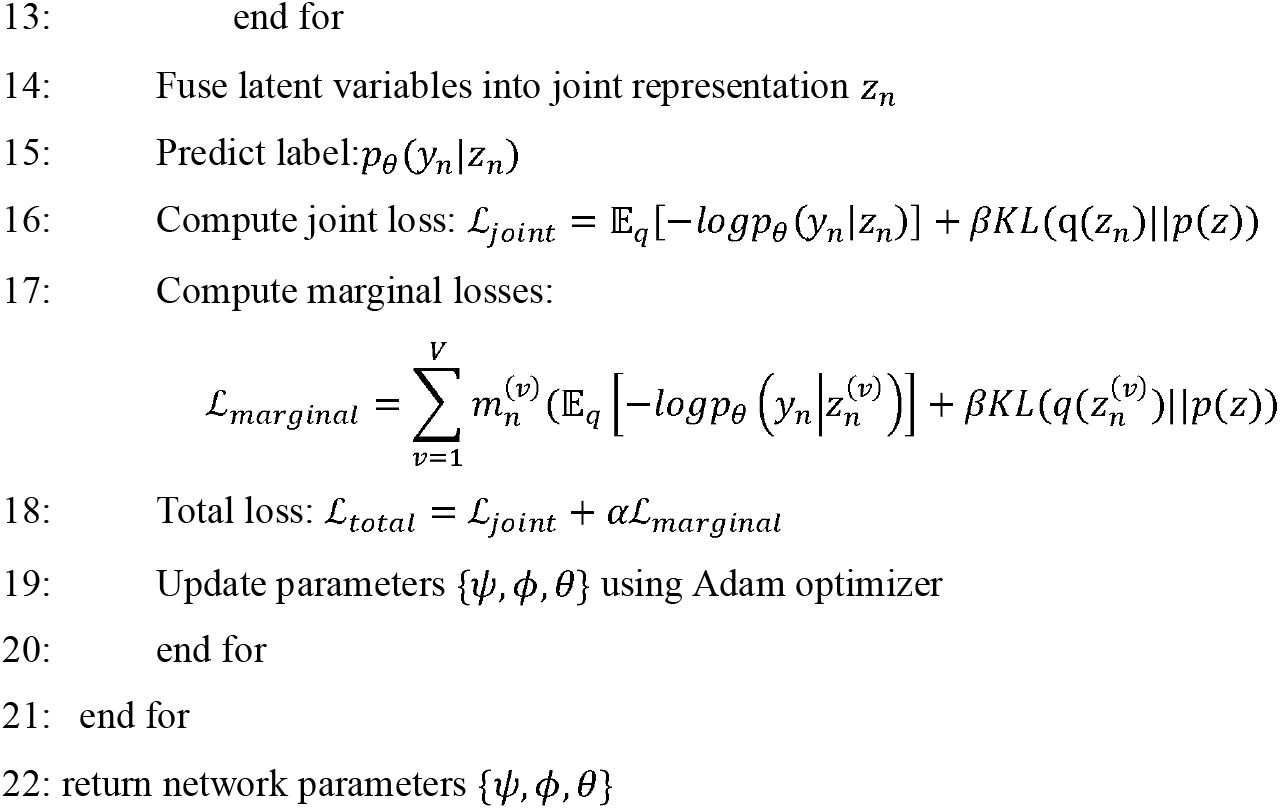

## 4. Experimental results and analysis

This section evaluates the effectiveness of ESSFS-IMO through comprehensive experiments. We first introduce the benchmark datasets and implementation details, then describe the compared baselines. Subsequently, we report quantitative results under varying missing-modality scenarios to assess predictive performance and robustness.

### 4.1. Benchmark datasets

The benchmark datasets utilized in this study were sourced from the Inflammatory Bowel Disease Multi-Omics Database (IBDMDB) (https://www.ibdmdb.org), a comprehensive public resource designed to facilitate the deep analysis of host-microbiome interactions in IBD. To align with our study goals and ensure a consistent basis for comparison with established literature, we focused on three primary omics modalities: metagenomics (mg), profiling the gut microbial community structure; metatranscriptomics (mt), characterizing the community-wide gene expression profile; and metabolomics (mb), capturing the repertoire of small molecule metabolites.

The initial dataset was characterized by its high dimensionality and heterogeneous missing pattern. Specifically, profile data were available for 1,638 samples at the metagenomic level (mg), 817 samples at the metatranscriptomic level (mt), and 546 samples at the metabolomic level (mb). To mitigate the challenges associated with high-dimensional data, such as noise and the risk of overfitting, a rigorous preprocessing pipeline was implemented for each omics type prior to analysis: (i) Metagenomics Data: Features that could not be accurately mapped to known microbial species were excluded from the analysis. This filtering step resulted in a curated set of 578 taxonomic features. A logarithmic transformation 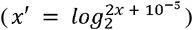 was subsequently applied to approximate a Gaussian distribution and reduce the influence of extreme outliers. (ii) Metabolomics Data: Only metabolites with confidently annotated compound identities were retained, yielding a refined dataset comprising 596 known metabolic features. (iii) Metatranscriptomics Data: Analysis was confined to features representing total metabolic pathways, resulting in a focused set of 421 functional features. Furthermore, to account for variations in scale and dynamic range across the different omics datasets, each feature within a specific omics type was normalized to a comparable scale using the operation: 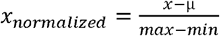, where μ is the mean value of the feature. This step is crucial for ensuring the stable and efficient training of neural network models. The final preprocessed dataset, with its curated features and normalized values, provides a robust foundation for evaluating multi-omics integration methods on IBD prediction.

### 4.2. Performance measures

The prediction accuracy (ACC) served as our primary performance evaluation index. To provide a comprehensive assessment of the compared methods, we also employed the F1 score and the area under the receiver operating characteristic curve (AUC) as additional evaluation metrics. Based on the confusion matrix (Table 1), which tabulates true positives (TP), false positives (FP), true negatives (TN), and false negatives (FN), these metrics are defined as follows: Accuracy (ACC) measures the overall proportion of correct predictions: 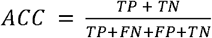 Score represents the harmonic mean of precision and recall: 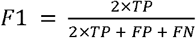. AUC is calculated by plotting the true positive rate 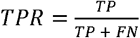 against the false positive rate 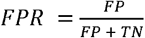 at various threshold settings and measuring the area under this curve. Higher values of ACC, F1 score, and AUC indicate better model performance.

**Table 1:** The number of samples included in different datasets.

| Dataset | Samples | IBD patients | Healthy |
| --- | --- | --- | --- |
| mb | 546 | 411 | 135 |
| mg | 1638 | 1209 | 429 |
| mt | 817 | 617 | 200 |
| all_mg+mb | 1796 | 1337 | 459 |
| all_mg+mt | 1638 | 1209 | 429 |
| all_mt+mb | 1064 | 815 | 249 |
| all_mg+mt+mb | 1796 | 1337 | 459 |
| common_mg+mb | 388 | 283 | 105 |
| common_mg+mt | 817 | 617 | 200 |
| commonmt+mb | 299 | 213 | 86 |
| common_mg+mt+mb | 299 | 213 | 86 |

### 4.3. Experimental settings

We used 5-fold cross-validation (CV) for unbiased evaluation. All samples were randomly split into five disjoint folds; the model was trained on four folds and tested on the remaining fold, iterating until each fold served as the test set once. This guarantees each sample is tested exactly once, yielding an unbiased estimate of generalization.

To maximize generalization, we performed an extensive hyper-parameter search. The key hyperparameters tuned included: the number of training epochs (tested within [10,000, 15,000, 20,000, 25,000]), the learning rate (explored within [0.0001, 0.001, 0.005, 0.01]), the dimensionality of the latent space (tested with [25, 50, 75]), and the dimensions and depth of the hidden layers in encoders and predictors (dimensions explored in [50, 100, 150, 200] with layer numbers ranging from 1 to 3). The weight balance parameter β was tested in the range of [0.001, 0.01, 0.1, 0.5, 1], while *α* was set to a constant value of 1.0. Through this process, the final learning rate was determined to be 0.005, the latent space dimension was set to 50, and the encoder and predictor hidden layer dimensions were set to 100 with 3 and 2 hidden layers, respectively. The weight balance parameter β was set to 0.001. All hyperparameters were tuned using grid search within each training fold, where 10% of the training data was further held out for validation. The final model was retrained on the entire training fold with the best hyperparameters and then evaluated on the corresponding test fold.

### 4.4. Compared methods

To thoroughly evaluate our model, we compared it against eight state-of-the-art baseline methods, which can be categorized as follows: (i) Traditional Machine Learning Methods: Support Vector Machine (SVM) [28] and Random Forest (RF) [29]: Two widely used classifiers for single-view data. For a fair comparison on multi-omics data, the features from all omics were simply concatenated into a single vector for input to these models. (ii) Unsupervised Multi-Omics Integration Methods: Generalized Canonical Correlation Analysis (GCCA) [30], Deep Generalized Canonical Correlation Analysis (DGCCA) [31], and Multi-Omics Factor Analysis v2 (MOFA+) [32]: These are popular unsupervised techniques for learning common latent representations from multi-view data. For these methods, the learned latent representations were first extracted and then used to train a Multilayer Perceptron (MLP) classifier for the downstream prediction task. (iii) Deep Learning-based Methods: OmiVAE [33]: An end-to-end deep learning model that combines a variational autoencoder with a classifier to derive latent representations from multi-omics data for sample classification. CPM-Nets [34]: A state-of-the-art method specifically designed for classifying incomplete multi-view data by learning a common latent representation. MOGONET [18]: A model for integrating multi-omics data through the construction of gene co-expression networks. For methods that require complete data (SVM, RF, GCCA, DGCCA, OmiVAE, MOGONET), mean imputation was applied to fill in the missing omics values prior to model training and evaluation.

### 4.5. Results and Discussions

This section evaluates predictive performance. To thoroughly assess its capability and identify the optimal model configuration, we first performed a systematic hyperparameter search, focusing particularly on the dimension *F* of the feature-selective layer—a key architectural component. A range of values from 10 to 150 was tested for *F* across all datasets. The overall trend in prediction accuracy is shown in Figure 2.

**Figure 2.**
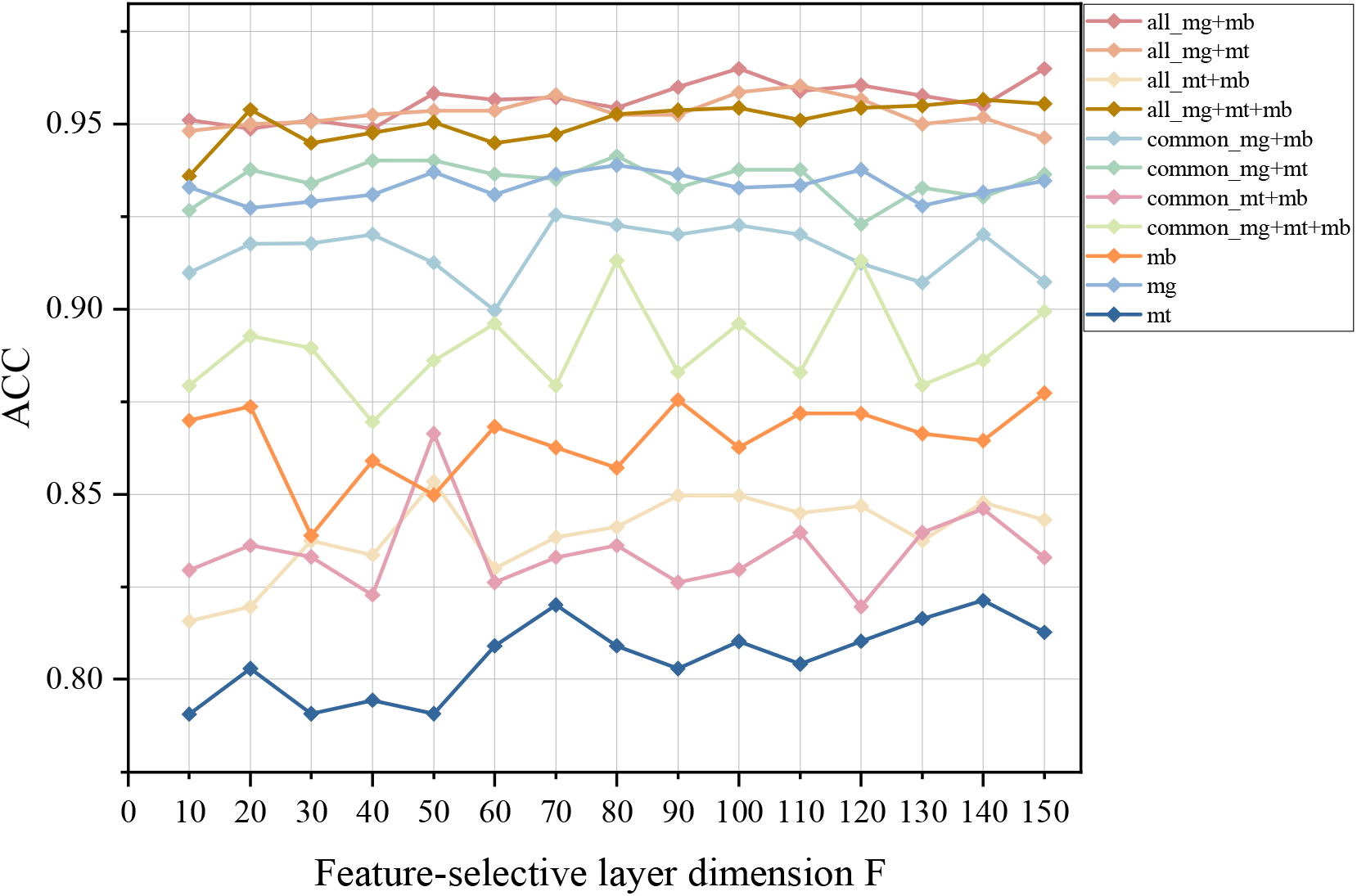
Experimental performance results of the ESSFS-IMO in terms of ACC with varying numbers of selected features (F) across different omics combinations. All results are based on 5-fold CV.

The results demonstrate that very small values of *F* generally lead to inferior performance, whereas larger settings tend to yield more stable and competitive outcomes. Specifically, the performance steadily improves as *F* increases up to around 100, beyond which the gain becomes marginal while the model complexity continues to rise. Considering both accuracy and computational efficiency, we fixed *F* = 100 for all subsequent experiments, as it consistently achieves near-optimal performance with lower variance across different datasets.

Tables 3–5 summarize the comparative results of various methods in disease prediction tasks, reporting three performance metrics: accuracy (ACC), F1 score, and area under the curve (AUC). ESSFS-IMO consistently outperforms all baseline methods across nearly all omics combinations and evaluation metrics, demonstrating its robustness and effectiveness in integrating incomplete multi-omics data for accurate disease prediction.

**Table 2:** Confusion matrix for prediction results.

|  | Reference Positive | Reference Negative |
| --- | --- | --- |
| Predicted Positive | True Positive (TP) | False Positive (FP) |
| Predicted Negative | False Negative (FN) | True Negative (TN) |

**Table 3:** Comparison of ESSFS-IMO with the Baseline Algorithm in Terms of ACC. All results are based on 5-fold CV.

|  | ESSFS-IMO | MOGONET | CPM-Nets | GCCA | DGCCA | MOFA | OmiVAE | RF | SVM |
| --- | --- | --- | --- | --- | --- | --- | --- | --- | --- |
| **all_mg+mb** | 0.964±0.006 | 0.888±0.012 | 0.839±0.015 | 0.735±0.018 | 0.802±0.016 | 0.763±0.022 | 0.862±0.017 | 0.890±0.011 | 0.772±0.024 |
| **all_mg+mt** | 0.958±0.008 | 0.919±0.010 | 0.872±0.012 | 0.877±0.014 | 0.832±0.016 | 0.731±0.027 | 0.918±0.010 | 0.902±0.012 | 0.785±0.023 |
| **all_mt+mb** | 0.850±0.029 | 0.772±0.028 | 0.578±0.037 | 0.752±0.032 | 0.748±0.031 | 0.738±0.030 | 0.673±0.034 | 0.755±0.033 | 0.742±0.035 |
| **all_mg+mt+mb** | 0.954±0.007 | 0.909±0.011 | 0.841±0.013 | 0.728±0.019 | 0.792±0.018 | 0.742±0.021 | 0.909±0.011 | 0.835±0.015 | 0.776±0.020 |
| **common_mg+mb** | 0.923±0.013 | 0.862±0.014 | 0.741±0.022 | 0.713±0.025 | 0.788±0.020 | 0.785±0.019 | 0.851±0.016 | 0.848±0.017 | 0.769±0.023 |
| **common_mg+mt** | 0.938±0.013 | 0.907±0.013 | 0.781±0.017 | 0.868±0.015 | 0.879±0.014 | 0.718±0.025 | 0.907±0.013 | 0.847±0.018 | 0.742±0.022 |
| **common_mt+mb** | 0.830±0.051 | 0.809±0.037 | 0.713±0.042 | 0.695±0.045 | 0.735±0.039 | 0.677±0.047 | 0.728±0.041 | 0.809±0.037 | 0.752±0.041 |
| **common_mg+mt+mb** | 0.896±0.392 | 0.825±0.029 | 0.752±0.032 | 0.732±0.034 | 0.785±0.027 | 0.695±0.040 | 0.831±0.026 | 0.825±0.029 | 0.754±0.035 |

**Table 4:** Comparison of ESSFS-IMO with the Baseline Algorithm in Terms of F1. All results are based on 5-fold CV.

|  | ESSFS-IMO | MOGONET | CPM-Nets | GCCA | DGCCA | MOFA | OmiVAE | RF | SVM |
| --- | --- | --- | --- | --- | --- | --- | --- | --- | --- |
| **all_mg+mb** | 0.976±0.005 | 0.948±0.007 | 0.785±0.016 | 0.837±0.015 | 0.891±0.011 | 0.879±0.012 | 0.915±0.010 | 0.948±0.007 | 0.888±0.013 |
| **all_mg+mt** | 0.972±0.005 | 0.960±0.006 | 0.808±0.013 | 0.905±0.009 | 0.925±0.008 | 0.818±0.016 | 0.942±0.007 | 0.960±0.006 | 0.885±0.013 |
| **all_mt+mb** | 0.903±0.018 | 0.882±0.018 | 0.635±0.031 | 0.879±0.019 | 0.862±0.021 | 0.825±0.023 | 0.832±0.022 | 0.882±0.018 | 0.858±0.021 |
| all_mg+mt+mb | 0.969±0.005 | 0.945±0.007 | 0.788±0.015 | 0.833±0.015 | 0.892±0.011 | 0.855±0.013 | 0.840±0.014 | 0.945±0.007 | 0.888±0.013 |
| common_mg+mb | 0.947±0.008 | 0.892±0.011 | 0.752±0.020 | 0.815±0.017 | 0.888±0.012 | 0.878±0.013 | 0.895±0.012 | 0.892±0.011 | 0.842±0.016 |
| common_mg+mt | 0.960±0.008 | 0.905±0.011 | 0.742±0.021 | 0.915±0.009 | 0.912±0.010 | 0.807±0.016 | 0.948±0.008 | 0.905±0.011 | 0.847±0.014 |
| common_mt+mb | 0.883±0.037 | 0.878±0.027 | 0.692±0.036 | 0.845±0.029 | 0.847±0.030 | 0.792±0.033 | 0.803±0.032 | 0.878±0.027 | 0.835±0.032 |
| common_mg+mt+mb | 0.929±0.026 | 0.878±0.021 | 0.741±0.027 | 0.801±0.024 | 0.838±0.022 | 0.848±0.021 | 0.718±0.031 | 0.878±0.021 | 0.835±0.025 |

**Table 5:** Comparison of ESSFS-IMO with the Baseline Algorithm in Terms of AUC. All results are based on 5-fold CV.

|  | ESSFS-IMO | MOGONET | CPM-Nets | GCCA | DGCCA | MOFA | OmiVAE | RF | SVM |
| --- | --- | --- | --- | --- | --- | --- | --- | --- | --- |
| all_mg+mb | 0.983±0.003 | 0.965±0.005 | 0.870±0.011 | 0.605±0.029 | 0.835±0.014 | 0.827±0.015 | 0.955±0.007 | 0.965±0.005 | 0.862±0.015 |
| all_mg+mt | 0.980±0.008 | 0.971±0.006 | 0.885±0.011 | 0.890±0.011 | 0.921±0.009 | 0.735±0.024 | 0.971±0.006 | 0.971±0.006 | 0.872±0.014 |
| all_mt+mb | 0.845±0.034 | 0.778±0.029 | 0.623±0.034 | 0.633±0.034 | 0.661±0.032 | 0.683±0.033 | 0.739±0.031 | 0.778±0.029 | 0.668±0.035 |
| all_mg+mt+mb | 0.970±0.011 | 0.963±0.007 | 0.874±0.011 | 0.637±0.027 | 0.819±0.014 | 0.770±0.017 | 0.968±0.006 | 0.963±0.007 | 0.862±0.015 |
| common_mg+mb | 0.950±0.042 | 0.922±0.019 | 0.868±0.018 | 0.625±0.029 | 0.898±0.012 | 0.856±0.014 | 0.931±0.011 | 0.922±0.019 | 0.775±0.024 |
| common_mg+mt | 0.970±0.012 | 0.933±0.013 | 0.853±0.016 | 0.922±0.010 | 0.875±0.013 | 0.695±0.024 | 0.922±0.012 | 0.933±0.013 | 0.755±0.021 |
| common_mt+mb | 0.860±0.040 | 0.871±0.026 | 0.633±0.034 | 0.543±0.039 | 0.829±0.019 | 0.681±0.034 | 0.741±0.031 | 0.871±0.026 | 0.770±0.029 |
| common_mg+mt+mb | 0.937±0.039 | 0.899±0.021 | 0.855±0.019 | 0.589±0.031 | 0.784±0.021 | 0.770±0.022 | 0.820±0.024 | 0.899±0.021 | 0.774±0.025 |

Notably, ESSFS-IMO achieves the highest average ACC of 0.964 on the all_mg+mb dataset, surpassing the second-best method (RF, ACC=0.890) by a significant margin of 7.4%. Similarly, on the all_mg+mt dataset, ESSFS-IMO attains an ACC of 0.958, which is 5.6% higher than the next best method (OmiVAE, ACC=0.918). These results clearly indicate that our method better leverages the complementary information from multiple omics types, even in the presence of missing data, to improve classification accuracy. Furthermore, when evaluated using the F1 score and AUC, ESSFS-IMO continues to exhibit superior performance. For instance, on the all_mg+mt dataset, ESSFS-IMO achieves an F1 score of 0.972 and an AUC of 0.980, both of which are the highest among all compared methods. This indicates that our model not only achieves high overall accuracy but also maintains a strong balance between precision and recall, along with excellent discriminatory power between classes. It is worth noting that while some methods perform competitively on certain datasets, they fail to consistently outperform ESSFS-IMO across all omics combinations. For example, on the more challenging all_mg+mt+mb datasets, which exhibit higher missing rates and inherent noise, ESSFS-IMO still achieves the best performance, underscoring its robustness in handling incomplete and complex multi-omics data. To further evaluate the robustness of ESSFS-IMO in handling incomplete data, we conducted experiments under various missing rates. we artificially constructed incomplete multi-omics datasets by randomly removing omics data for each sample. The missing rate R for a dataset is defined as the proportion of missing omics instances relative to the total number of potential omics instances across all samples, calculated as:

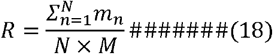

where *N* is the total number of samples, *M* is the total number of omics types (3 in this study), and *m*_*n*_ denotes the number of missing omics types for the *n*-th sample. It is ensured that each sample retains at least one omics type. We conducted extensive experiments under varying missing rates ranging from 0.1 to 0.9.

The accuracy results across four different omics combinations are summarized in Table 6 and visualized in Figure 3. Overall, across all omics combinations, the model demonstrates robust performance as the missing rate increases. And while a gradual downward trend in performance is observable—a natural consequence of having less information available, but the decline is notably graceful and non-catastrophic. For instance, the median accuracy for most datasets remains well above 0.85 until a missing rate of 0.4-0.5, and retains clinically or biologically relevant predictive power even under extreme missingness conditions of 0.8-0.9. This underscores the model’s core capability to extract and integrate meaningful patterns from the available data, rather than being crippled by what is absent. This observation highlights the effectiveness of our joint-omics encoder and the information bottleneck-based training in mitigating the adverse effects of missing data, confirming the model’s robustness in real-world scenarios where data completeness is rare.

**Table 6:**
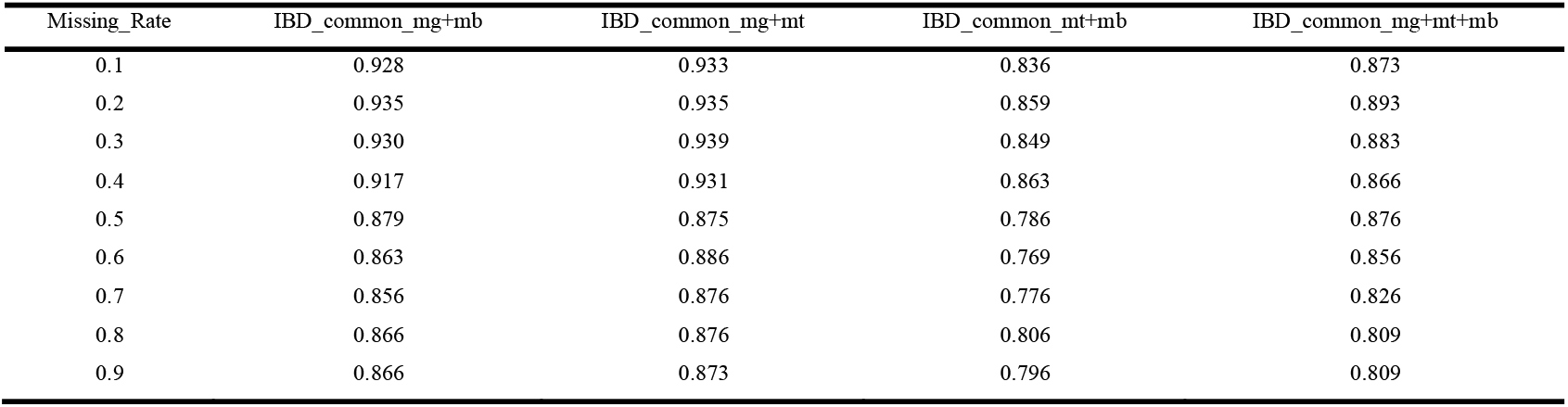
Experimental performance results of the ESSFS-IMO in terms of ACC on “common” datasets with different missing rate. All results are based on 5-fold CV.

| Missing_Rate | IBD_common_mg+mb | IBD_common_mg+mt | IBD_common_mt+mb | IBD_common_mg+mt+mb |
| --- | --- | --- | --- | --- |
| 0.1 | 0.928 | 0.933 | 0.836 | 0.873 |
| 0.2 | 0.935 | 0.935 | 0.859 | 0.893 |
| 0.3 | 0.930 | 0.939 | 0.849 | 0.883 |
| 0.4 | 0.917 | 0.931 | 0.863 | 0.866 |
| 0.5 | 0.879 | 0.875 | 0.786 | 0.876 |
| 0.6 | 0.863 | 0.886 | 0.769 | 0.856 |
| 0.7 | 0.856 | 0.876 | 0.776 | 0.826 |
| 0.8 | 0.866 | 0.876 | 0.806 | 0.809 |
| 0.9 | 0.866 | 0.873 | 0.796 | 0.809 |

**Figure 3.**
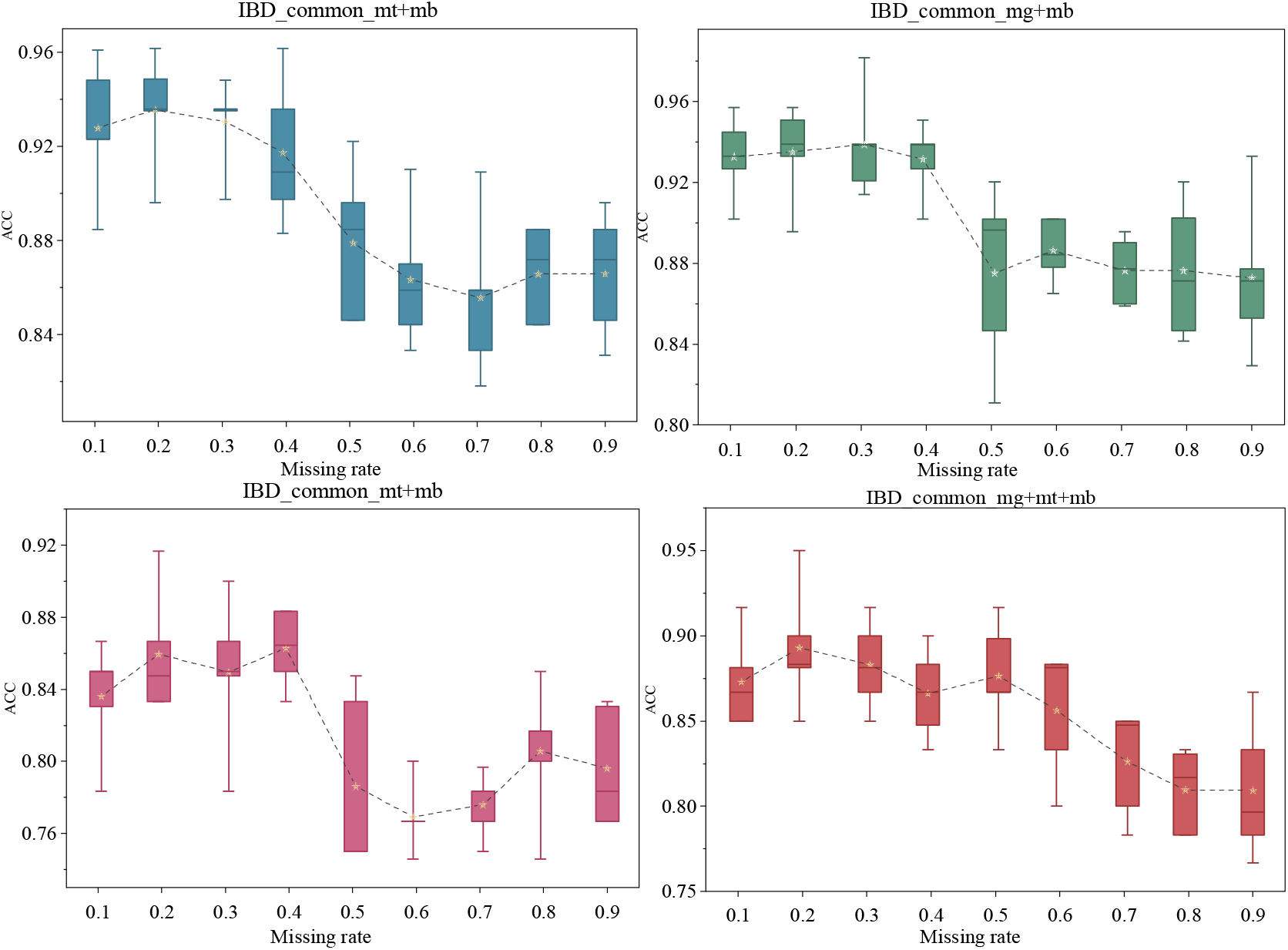
Model performance under different missing rates on “common” sample subsets. The star represents the average value. All results are based on 5-fold CV.

Having established the robustness of our model under various missing conditions, we next validate the effectiveness of its core integrative capability. Specifically, we investigate whether this robust framework successfully translates into a tangible performance gain by leveraging complementary information from multiple omics sources, as compared to relying on a single omics type.

As shown in Figure 4, multi-omics integration consistently yields superior predictive performance over any single-omics approach. Notably, the model achieves its peak accuracy (0.969) on the IBD_all_mg+mb dataset significantly outperforming the results obtained from using metabolomics alone (mb, 0.845-0.927) and metagenomics alone (mg, 0.948-0.982). This indicates that our integration mechanism does not merely compensate for missing views but actively synthesizes novel and discriminative information from the interplay between different omics layers. Furthermore, the three-omics combination IBD_all_mg+mt+mb also achieves commendable performance (accuracy up to 0.964), reinforcing the capability of our model to harness the complexity of multiple data sources without being overwhelmed. It is particularly noteworthy that even in combinations involving the relatively weaker single-omics performer metatranscriptomics (mt, 0.76-0.83), its integration with other omics types still leads to a substantial performance boost. This underscores a key advantage of our method: the ability to extract and amplify the valuable signal contained within each individual omics type, resulting in a final prediction that is greater than the sum of its parts. In summary, these results conclusively demonstrate that the superior performance of our model is fundamentally driven by its effective and powerful multi-omics data integration capacity.

**Figure 4.**
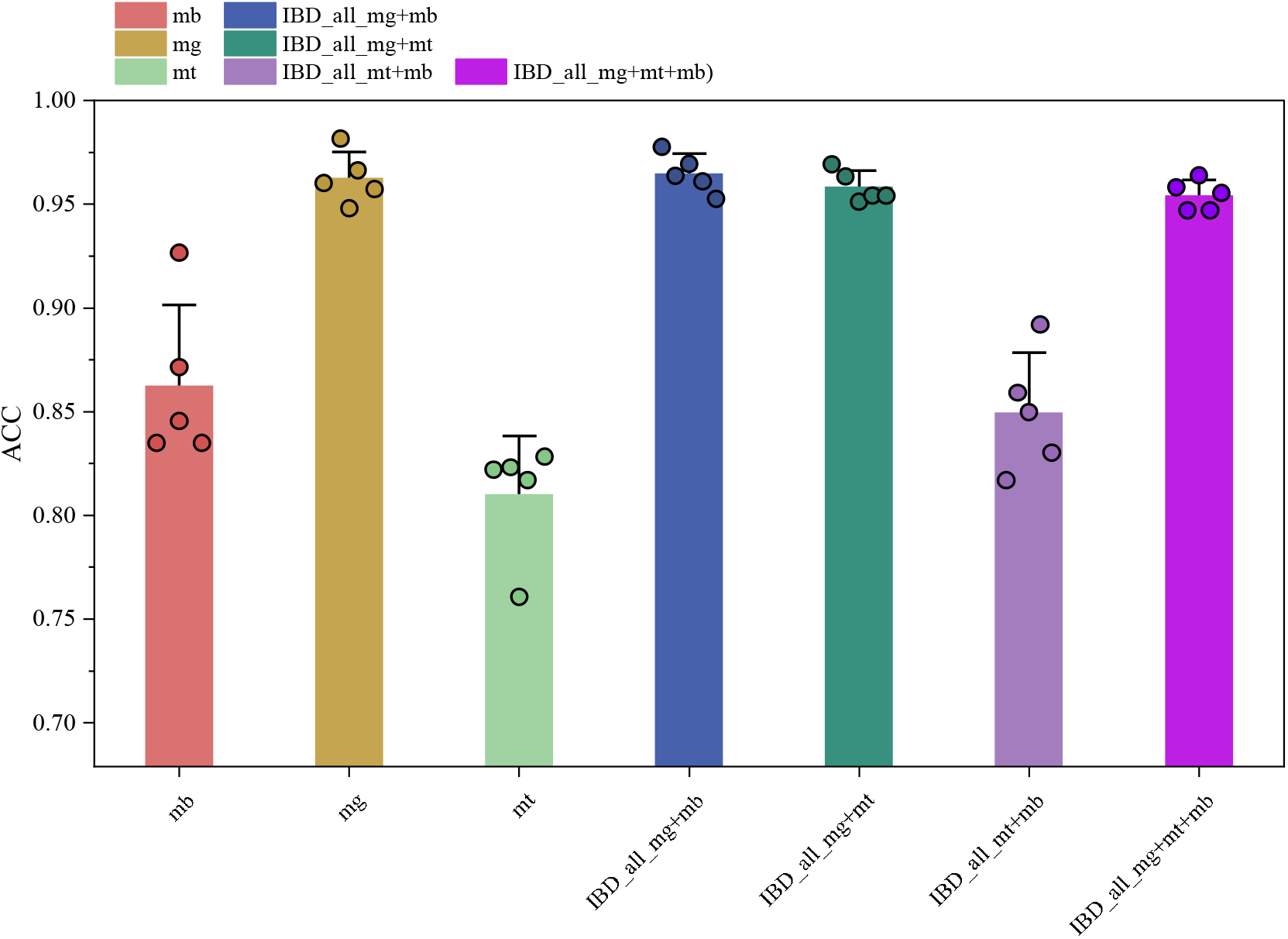
Performance comparison between multi-omics and single-omics models. The error bar represents plus one standard deviation. All results are based on 5-fold CV.

The superior performance achieved through multi-omics integration, as evidenced in Figure 4, establishes the core effectiveness of our model. To further demonstrate its unique capability to extract value from incomplete samples, we directly compared its performance on the full dataset (all) against the complete-case subset (common). This comparison, detailed in Figure 5, serves to quantify the benefit of utilizing all available data, including samples with missing views.

**Figure 5.**
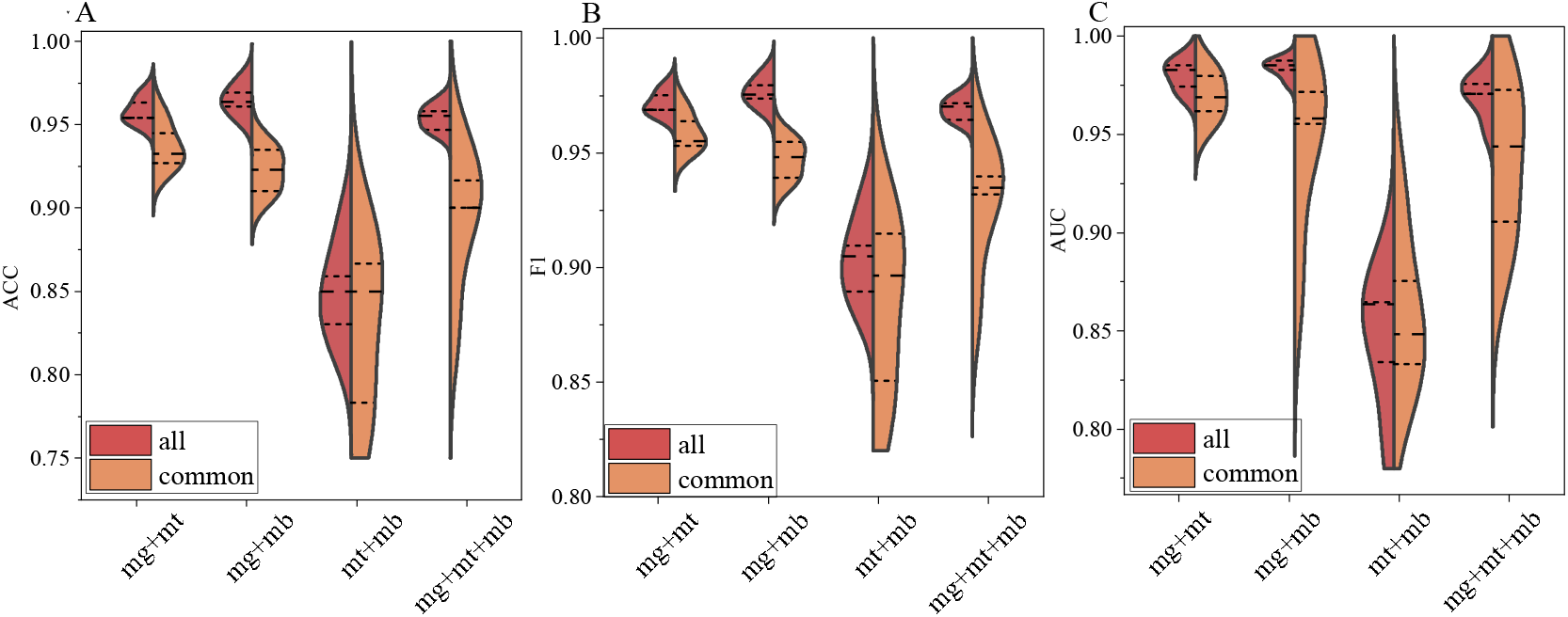
Performance comparison between models trained on “all” samples and on “common” samples. All results are based on 5-fold CV.

The results unequivocally show that our model consistently achieve better results on the “all” datasets compared to their “common” counterparts across all omics combinations. For instance, a significant performance gain is observed on the mg+mt+mb combination. Here, the model achieves a mean accuracy of 0.954 on the “all” dataset, a substantial improvement over the 0.896 attained on the far smaller “common” dataset, and this performance difference was consistently reflected in all key indicators: the F1 score increased from 0.929 to 0.969, and the AUC rose from 0.937 to 0.970.It demonstrates that our framework not only compensates for missingness but capitalizes on the increased informational breadth available in the larger cohort to synthesize a more robust and generalizable representation. This pattern is also clearly evident in other combinations, such as mg+mt. Rather than being hampered by missing data, the model successfully converts the partial information from a broader cohort into a definitive net performance increase.

## 5. Conclusions

In this work we present ESSFS-IMO, a robust framework for incomplete multi-omics integration that incorporates sample-specific feature selection, entropy-guided optimization and variational information bottleneck learning. First, a Dynamic Gamma network with a Concrete distribution–based selector are employed to adaptively identify crucial features for per sample, reducing redundancy in high-dimensional omics. Second, an entropy-based annealing strategy dynamically controls the feature selection sharpness, allowing the model to balance exploration and exploitation during training. Third, a variational backbone with variance-weighted fusion integrates the selected features from different modalities, yielding compact and discriminative latent representations under arbitrary missing patterns.

To evaluate our approach, we conducted extensive experiments on IBD multi-omics datasets. The results demonstrate that ESSFS-IMO achieves superior predictive performance compared with several state-of-the-art methods, particularly in scenarios with high missing rates.

Although this study mainly focuses on IBD cohorts, the proposed framework is broadly applicable to other biomedical tasks involving incomplete and heterogeneous multi-omics data. In the future, we aim to extend ESSFS-IMO to longitudinal and survival analysis, and to incorporate pathway- or graph-level biological priors to enhance interpretability and clinical relevance.

## Data Availability

All data produced are available online at https://www.ibdmdb.org

## Acknowledgments

Research on this work is partially supported by grants from the National Natural Science Foundation of China (No. 62566041).

